# Post-COVID-19 Memory Complaints: Prevalence and Associated Factors

**DOI:** 10.1101/2022.01.23.22269525

**Authors:** Mashrur Ahmed, Simanta Roy, Sreshtha Chowdhury, Mohammad Azmain Iktidar, Sharmin Akhter, A. M. Khairul Islam, Mohammad Delwer Hossain Hawlader

**Author notes:** These authors contributed equally (Mashrur Ahmed, Mohammad Azmain Iktidar).

## Abstract

**Introduction:** Memory complaints resulting from COVID-19 may have a significant impact on the survivors’ quality of life. Unfortunately, there is insufficient information available on memory loss and its relationship to COVID-19. Therefore, the purpose of this research was to determine the prevalence of memory complaints in post-COVID-19 patients and to find potential contributing factors.

**Method:** A cross-sectional survey was conducted on 401 individuals who had previously been diagnosed with COVID-19 at four COVID testing centers situated across Bangladesh. The MAC-Q questionnaire was used to evaluate memory. A binary logistic regression model was fit to study the variables related to memory complaints, with a p-value of <0.05 deemed statistically significant.

**Result:** Memory complaints was prevalent in 19.2% of the post-COVID patients. Individual predictor analysis revealed that among the treatment modalities, steroids and antibiotics were associated with impaired memory. Multiple logistic regression showed that individuals who recovered from COVID-19 within six to twelve months were more likely to have memory deficits. Even though age, sex, oxygen demand, and hospitalization were not linked with memory complaints, rural residents exhibited more significant memory complaints than urban residents.

**Conclusion:** Nearly one-fifth of the COVID-19 patients suffer from various degrees of memory complaints within one year. However, no association was found between COVID-19 severity to memory complaints.

## Introduction

The ongoing COVID-19 pandemic, caused by the recently identified severe acute respiratory syndrome coronavirus-2 (SARS-CoV-2), has taken the world off guard, with millions of people worldwide currently dealing with its severe consequences. Although COVID-19 primarily affects the respiratory system, it is increasingly recognized as a systemic disease.^1^ Our previous experience with members of the same coronavirus family (SARS and MERS), which caused two significant epidemics of much lower magnitude in the past, has taught us that the severe consequence of such outbreaks is not limited to acute problems alone. Following these infections, long-term neuro-metabolic and neuropsychiatric effects have been observed.^2^

Recent researches have demonstrated that COVID-19 can involve the nervous system.^3–5^ Lu et al. (2020) discovered that severe COVID-19 patients might lose grey matter volume in the brain’s frontal lobe.^6^ Grey matter reduction following COVID-19 is also reported by Douaud et al. (2021).^7^ As grey matter aids in processing information in the brain, allowing people to manage their movements, memories, and emotions, grey matter deficiency can result in significant brain problems and difficulties.^8^ SARS-CoV-2 has been reported to invade peripheral olfactory neurons ^10^ which allows for trans-synaptic viral spread to cortical regions, including the hippocampus.^11^ Since the hippocampus in humans plays a pivotal role in episodic memory and spatial memory ^12,13^, hippocampal damage may lead to memory impairment. This direct impact on the hippocampus can be amplified by COVID-19 associated immune response that compromises the blood-brain barrier ^14^ and disseminated intravascular coagulation (DIC) related vascular dysfunction.^15^

Brain in humans represents only 2% of total body weight; however, at rest, it consumes 20% of the body’s total basal oxygen.^16^ This high oxygen demand makes the brain, especially the hippocampus, very vulnerable to hypoxia.^17^ It is well documented that COVID-19 causes hypoxia via multiple pathophysiologic processes.^18^ Moreover, critical care and medical oxygen crises worldwide, especially in low-and middle-income countries (LMICs) during this pandemic, have led to preventable anoxic and hypoxic brain injuries.^19^ Furthermore, statewide lockdown in many countries has often hindered and delayed people’s access to health care, contributing to irreversible anoxic and hypoxic damage to the brain.^20^ A small study conducted on 32 COVID-19 patients found 23 of them having memory complaints.^21^ Elevated glucocorticoid concentrations have been seen to be correlated with decreased hippocampal volume.^22^ As the hippocampus is one of the essential neuroanatomic substrates for memory, glucocorticoid-induced changes in hippocampal structure and function may disrupt memory processes that rely on it.^23^ Since steroid use in treating COVID-19 is prevalent because it can reduce mortality and morbidity ^24,25^, it can also cause memory complaints.

Arentz et al. (2020) found, more than two-thirds of COVID-19 patients admitted to intensive care units required mechanical ventilation, and all of them developed acute respiratory distress syndrome (ARDS) within three days.^26^ In addition, several studies observing the long-term outcome of patients who have suffered from ARDS have found loss in memory function ^27,28^, even lasting up to five years.^29^

As of October 2021, over 234 million people have recovered from COVID-19 ^30^; however, various symptoms have been observed to persist even after recovery from this disease.^31^ A recent study on non-hospitalized COVID-19 patients observed that 11% reported memory problems eight months after recovery.^32^ Apart from these, post-COVID-19 memory complaints is still an unexplored area of research. Therefore, it is crucial to estimate the prevalence of memory complaints following COVID-19 recovery and investigate its associated factors.

## Methods

### Study Design & Duration

This cross-sectional study was conducted from August to October 2021.

### Study Site & Participants

A total of 850 individuals were selected by systematic sampling from the patient records of four randomly selected COVID testing sites located across Bangladesh. Telephone numbers of all these individuals were collected from the records and they were contacted via telephone to enter the study; among which 306 individuals didn’t receive the phone call, 32 individuals had already died, 68 individuals didn’t meet the inclusion criteria, 39 individuals refused to participate and 4 individuals incompletely filled up the questionnaire. Therefore, the research included a sample of 401 post-COVID-19 patients (Figure 1). The inclusion criteria were: (1) lack of pre-existing neurological or mental disorders (i.e., dementia, history of stroke, Alzheimer’s, etc.), (2) the absence of depression, (3) the absence of use of benzodiazepines, antidepressants, or neuroleptics, (4) individuals who suffered from COVID-19 between 3 and 12 months before, and (5) providing informed consent.

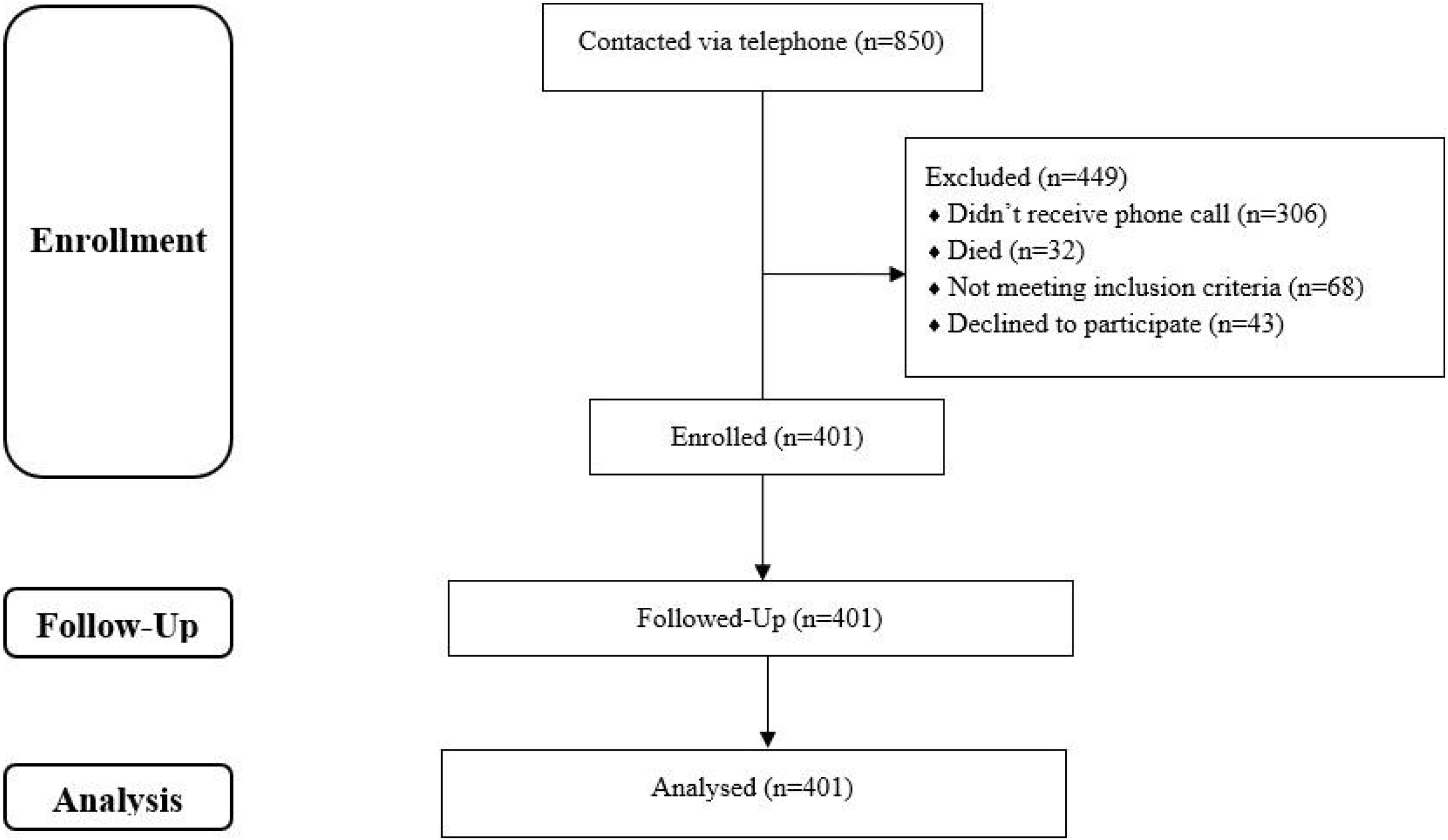

### Data collection tools

Before data collection, a semi-structured questionnaire was developed. This questionnaire has two parts. 1) Demographics and COVID-19 related history 2) Memory Complaint Questionnaire (MAC-Q). In the first part, demographic characteristics (Age, Sex, and Living region) and COVID-19 related clinical history (COVID-19 severity, oxygen requirement, hospitalization, treatments taken, co-morbidities, and duration since recovery) were collected. COVID-19 severity was assessed using a recently developed scoring system.^33^

The second part consists of the MAC-Q ^34^, a systematically widely validated self-report memory questionnaire. This questionnaire was developed to evaluate memory loss associated with aging. It consists of six questions on memory functioning in daily circumstances (e.g., remembering a phone number they use at least once a week). The subject is asked to compare and evaluate their current performance to that of their younger self. The MAC-Q total score ranges from 7 to 35, with higher values indicating subjective memory loss. Scores higher than or equal to 25 are indicative of memory decline. As a result, the participants in this research were split into two groups: those without significant memory complaints (MAC-Q scores of <25) and those with significant memory complaints (MAC-Q scores of ≥25) as suggested by Crook et al. (1992).^34^

The entire questionnaire was then inputted into Google Forms without any randomization of items for online distribution and tested for usability and technical functionality. The form had fifteen questions distributed over two pages. Mandatory items were highlighted with a red asterisk and relevant non-response option was present. Respondents were able to review their answers through the back button and change their response if they deemed necessary. The survey never displayed a second time once the user had filled it in to prevent duplicate entries.

### Data collection

Individuals who suffered from COVID-19 between 3 and 12 months before participants were contacted by trained volunteers via telephone and described the research in detail. Once the individuals were ascertained of meeting the inclusion criteria and consented to voluntary participation in the study, link to a web-based survey created by Google Forms was sent via SMS making it a closed survey. The survey wasn’t announced or advertised anywhere else. Of the 405 eligible participants who agreed to participate 401 participants completed the entire questionnaire (completion rate: 99.01%), incomplete questionnaires were excluded from analysis.

### Ethics

The Institutional Review Board of North South University approved the research (Approval no 2021/OR-NSU/IRB/0402), and all participants provided informed consent. Wherever feasible, the 1964 Declaration of Helsinki and later modifications and comparable ethical standards were followed. Data collection was voluntary and no incentives was offered to participants. Data were only accessible to the authors and was not disclosed anywhere. All the reporting was done according to the Checklist for Reporting Results of Internet E-Surveys (CHERRIES) guidelines.^35^

### Statistical analysis

The data has been processed and analyzed using the Stata version 16.0. Categorical data were summarized using frequency and relative frequency. For MAC-Q score, total score was collapsed into a binary variable of 0 (MAC-Q scores of <25) and 1 (MAC-Q scores of ≥25) and then a binary logistic regression model was fit to investigate the factors associated with memory complaints. A p-value of <0.05 was considered statistically significant.

## Result

Table 1 depicts the baseline characteristics of the study participants. Among the 401 participants, the majority were between 30-52 years of age (51.87%), male (64.84%), and from urban areas (89.03%). Most participants suffered from mild to moderate disease (56.11%), whereas 23.94% and 19.95% suffered from severe and critical diseases. Nearly half of the participants required hospitalization, and 39.40% required oxygen support. In addition, 52.87% of participants received antibiotics, 49.63% received steroids, 28.18% received anticoagulants, 15.46% received remedesevir, and 1.25% received tocilizumab. Hypertension (22.94%) and diabetes (21.95%) were the two most common. Two-thirds of participants recovered from COVID-19 6 to 12 months before, and the rest recovered 3 to 6 months back. 19.2% of the respondents suffered from memory complaints.

**Table 1:**
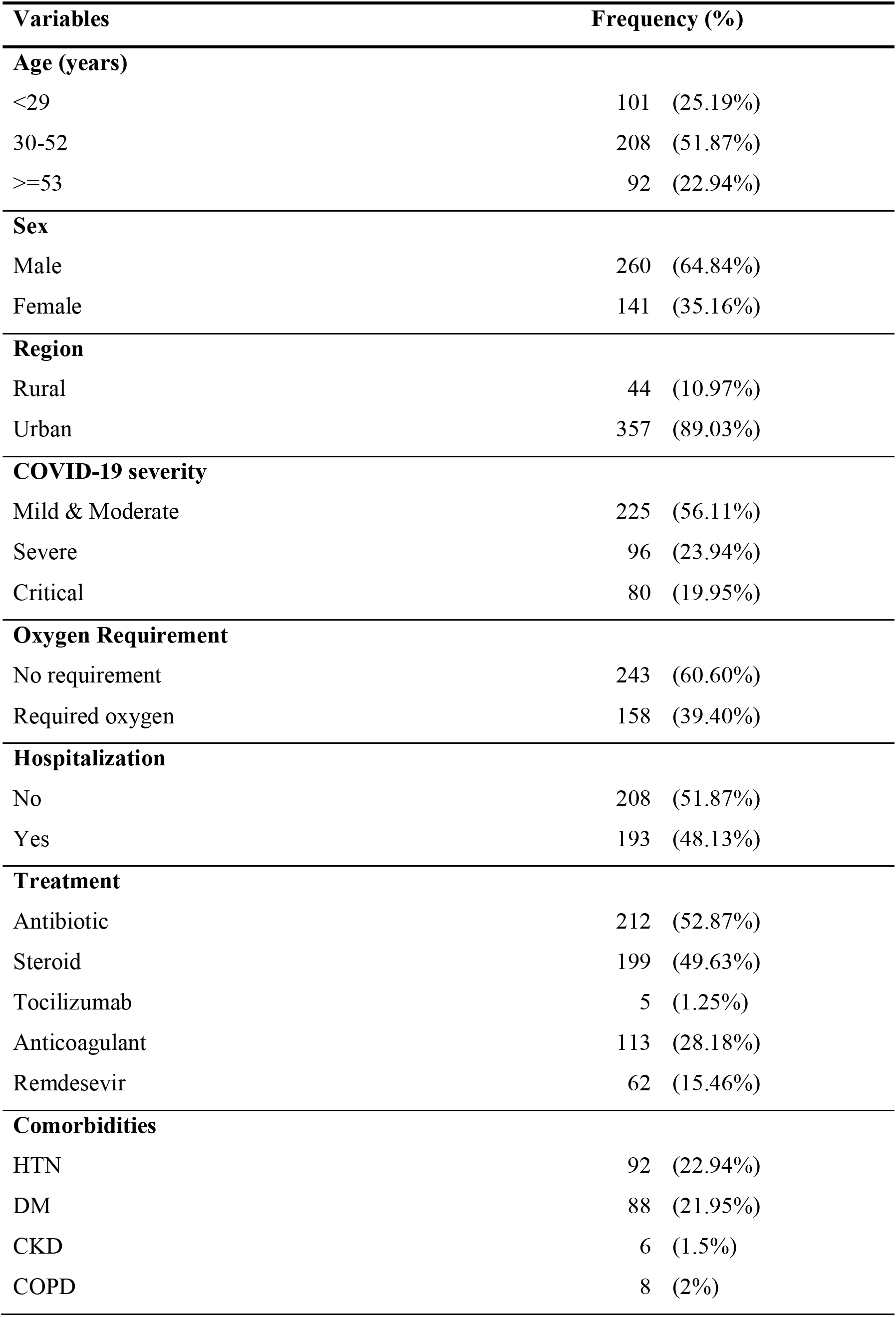
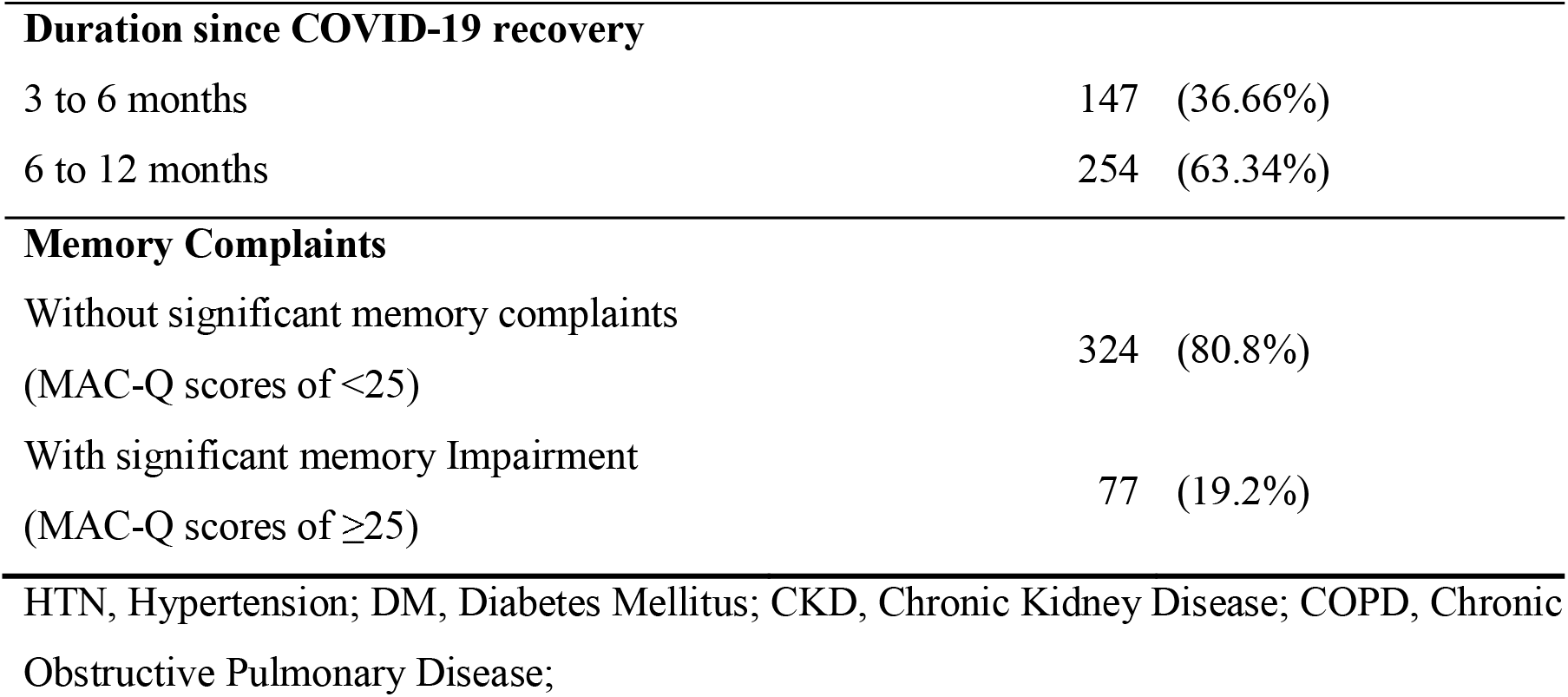
Baseline characteristics of study participants (n=401)

| **Variables** | **Frequency (%)** |
| --- | --- |
| **Age (years)** |  |
| <29 | 101 (25.19%) |
| 30-52 | 208 (51.87%) |
| >=53 | 92 (22.94%) |
| **Sex** |  |
| Male | 260 (64.84%) |
| Female | 141 (35.16%) |
| **Region** |  |
| Rural | 44 (10.97%) |
| Urban | 357 (89.03%) |
| **COVID-19 severity** |  |
| Mild & Moderate | 225 (56.11%) |
| Severe | 96 (23.94%) |
| Critical | 80 (19.95%) |
| **Oxygen Requirement** |  |
| No requirement | 243 (60.60%) |
| Required oxygen | 158 (39.40%) |
| **Hospitalization** |  |
| No | 208 (51.87%) |
| Yes | 193 (48.13%) |
| **Treatment** |  |
| Antibiotic | 212 (52.87%) |
| Steroid | 199 (49.63%) |
| Tocilizumab | 5 (1.25%) |
| Anticoagulant | 113 (28.18%) |
| Remdesevir | 62 (15.46%) |
| **Comorbidities** |  |
| HTN | 92 (22.94%) |
| DM | 88 (21.95%) |
| CKD | 6 (1.5%) |
| COPD | 8 (2%) |

|  |  |  |
| --- | --- | --- |
| **Duration since COVID-19 recovery** |  |  |
| 3 to 6 months | 147 | (36.66%) |
| 6 to 12 months | 254 | (63.34%) |
| **Memory Complaints** |  |  |
| Without significant memory complaints<br>(MAC-Q scores of <25) | 324 | (80.8%) |
| With significant memory Impairment<br>(MAC-Q scores of $\geq 25$) | 77 | (19.2%) |
| HTN, Hypertension; DM, Diabetes Mellitus; CKD, Chronic Kidney Disease; COPD, Chronic Obstructive Pulmonary Disease; |  |  |

Unadjusted and adjusted results of binary logistic regression of study variables are presented in Table 2. The unadjusted results show the region, duration since recovery from COVID-19, and co-existing respiratory disease to be significantly associated with post-COVID-19 memory complaints. Among these factors, only region and duration since recovery from COVID-19 retained their significance after adjustment. Rural people were 3.6 times more likely to suffer from post-COVID-19 memory complaints than urban people (AOR=3.63, 95%CI: 1.72 to 7.67, p=0.001). Also, memory complaints tended to become more prevalent as longer time progressed after COVID-19 recovery. People who suffered from COVID-19 6 to 12 months ago had a higher tendency to develop memory complaints than people who suffered from COVID-19 3 to 6 months ago (AOR=6.56, 95%CI: 1.16 to 38.18, p=0.033). Participants receiving steroids and antibiotics as a part of their COVID-19 treatment were 65% and 51% more likely to develop memory complaints, respectively. Patients with co-existing chronic obstructive pulmonary disease (COPD) are almost three times more likely to suffer from memory complaints. However, it loses its significance after adjusting for other factors. No association was found between age, Sex, COVID-19 severity with post-COVID-19 memory complaints.

**Table 2:**
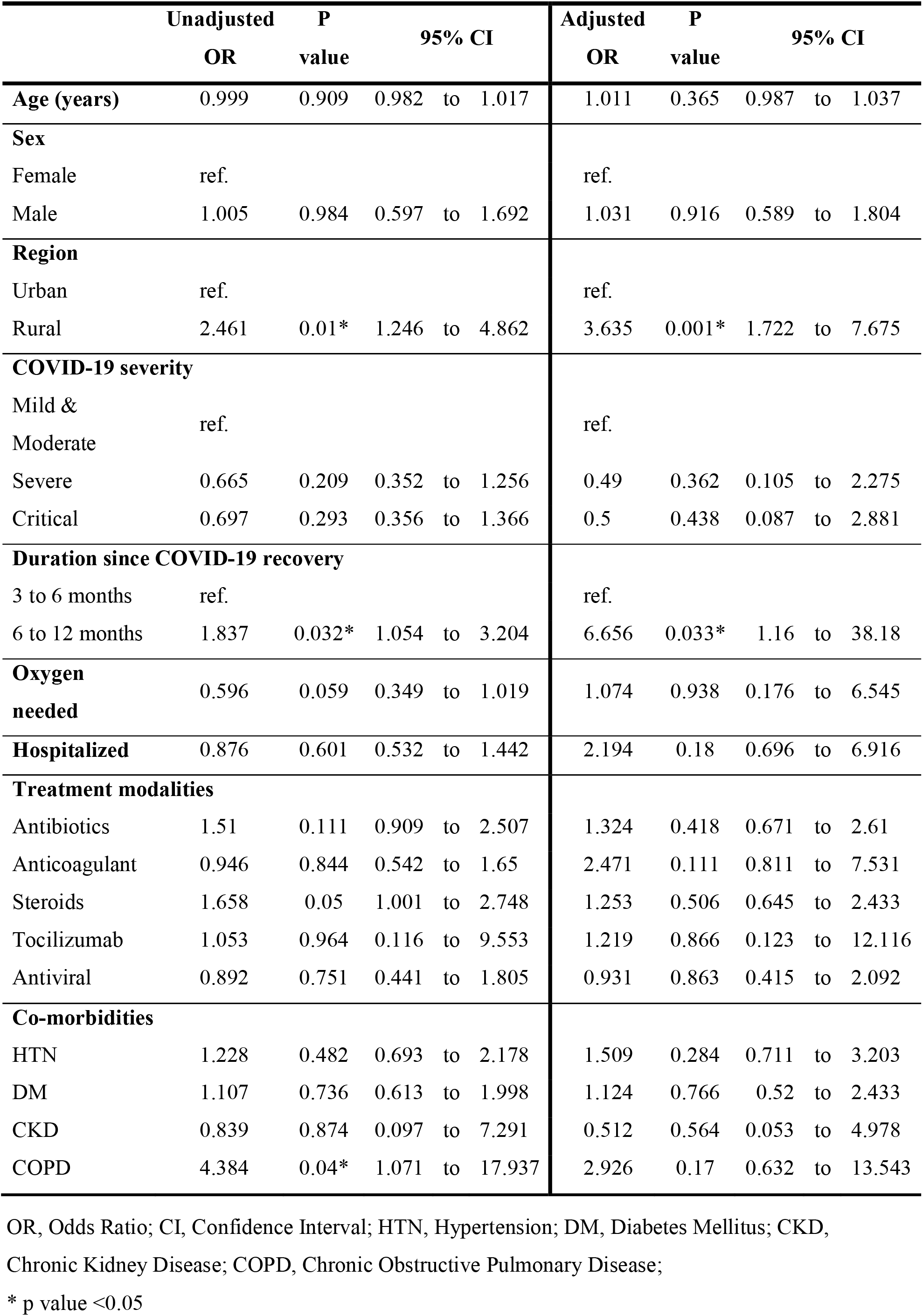
Associated factors of memory complaints in study participants (n=401)

## Discussion

Our results reveal that memory complaints is prevalent among one-fifth of the patients within one year of the post-COVID-19 recovery period. In contrast, Søraas et al. (2021) reported an 11% prevalence eight months after COVID-19 recovery.^32^ Our study included COVID-19 patients of all severity from mild to critical; on the other hand, Søraas et al. (2021) included only relatively mild and non-hospitalized cases, which may explain their lower prevalence.^32^ Our study also revealed that memory complaints was more likely to happen as time passed from COVID-19 recovery. For example, participants who recovered 6 to 12 months back were 6.6 times more likely to have memory complaints than the people who recovered 3 to 6 months ago (AOR=6.56, 95%CI: 1.16 to 38.18, p=0.033).

A cross-sectional survey among 1520 people of Bangladesh revealed that rural residents had a poor level of knowledge and practice associated with COVID-19.^36^ Moreover, many rural people rely on traditional healers for treatment purposes which may hamper their receipt of evidence-based treatment.^37^ Furthermore, healthcare facilities are also scarce in rural regions.^38^ These factors may contribute to the higher likelihood of memory complaints of the rural people than urban dwellers (AOR=3.63, 95%CI: 1.72 to 7.67, p=0.001).

Pre-existing COPD was significantly associated with post-COVID-19 memory complaints in bivariate analysis of our study, although it didn’t retain significance in multivariate analysis, COPD patients were three times more likely to have memory complaints than individuals. This finding is consistent with Huppert (1982), who reported chronic hypoxia due to COPD may lead to memory complaints.^39^

Steroid and antibiotic therapy participants showed a somewhat increased chance of experiencing memory complaints than other treatment groups. American Academy of Neurology suggests antibiotics may be more closely associated with delirium and other brain problems than previously thought.^40^ According to the available research, long-term glucocorticoid exposure may impair cognition by altering hippocampal function and architecture progressively and irreversibly. However, it is unclear if these acute hormonal effects on memory retrieval processes contribute to the problems observed with chronic exposure to glucocorticoids related to memory deficiencies.

Previous studies suggested that COVID-19 infection severity affected cognitive disorders.^41^ However, our study found no significant relation of memory complaints with COVID-19 severity.

One of the drawbacks of this study is the lack of a control group or a similar cohort study that may provide a better picture of the problem. Generalizability is also an issue as the sample size is comparatively smaller than the population size of COVID-19 recovered patients. This study also did not administer any neuropsychological test to assess memory complaints. Therefore, a future longitudinal study with a larger sample adminstering neuropsychological tests and with a control group is warranted to understand the condition better.

## Conclusion

One-fifth of the COVID-19 patients develop memory complaints within a year. The reason behind this memory complaints is still unclear. Since the quality of life of an individual is directly related to memory complaints ^41^, further investigations should be done to evaluate the etiology of post-COVID-19 memory complaints and to take necessary actions to minimize its prevalence.

## Data Availability

All data produced in the present work are contained in the manuscript

## Acknowledgment

None

